# Cohabitation timing and childbearing or current pregnancy among married adolescents in Bangladesh: a complex-survey analysis of the 2022 BDHS

**DOI:** 10.64898/2026.07.21.26358612

**Authors:** Md. Ikramul Haq Khan, Israt Jahan, Mohammad Amirul Islam, Rohan Dalpatadu, Mohammad Karim

## Abstract

**Objectives:** To describe the prevalence of having had a live birth or being currently pregnant among ever-married adolescents in Bangladesh and examine its associations with cohabitation timing and partner characteristics among currently married adolescents.

**Design:** Cross-sectional secondary analysis of the 2022 Bangladesh Demographic and Health Survey (BDHS).

**Methods:** Descriptive estimates used 2,449 ever-married respondents aged 15–19. The long questionnaire was administered to 1,635; 1,603 were currently married, of whom 1,601 had complete information for the regression. Weighted logistic regression included current age, respondent education, five wealth quintiles, cohabitation age, husband education, signed spousal age gap, residence and division. Sampling weights, strata and primary sampling units were incorporated using a stratified cluster-based Taylor variance estimator. Current working status was evaluated in a sensitivity model. An asset-based principal component was examined separately as an exploratory sensitivity analysis.

**Results:** Among ever-married adolescents, weighted outcome prevalence was 59.7% (95% confidence interval [CI] 57.6–61.7). In the expanded model, cohabitation at ages 15–17 and 18–19, relative to below 15, was associated with lower odds (adjusted odds ratios [AORs] 0.170, 95% CI 0.113–0.256; and 0.026, 0.015–0.045). A signed spousal age gap of at least 11 years, versus at most five years, was associated with higher odds (AOR 1.752, 1.241–2.474). The higher-husband-education comparison was inconclusive (AOR 0.612, 0.338–1.109). Estimates for cohabitation timing changed substantially after age adjustment. Mean held-out-PSU AUC was 0.793 and calibration slope was 0.885. An exploratory specification diagnostic indicated possible misspecification (*p ≈* 0.0205).

**Conclusions:** Cohabitation timing and spousal age gap were associated with childbearing or current pregnancy in the analysed population. Differences in time available for childbearing, selection into marriage, and interview-time measurement of covariates constrain interpretation. These findings do not estimate the effects of delaying marriage or implementing a policy. Regression estimates remain provisional because long-questionnaire weighting requires confirmation.

## 1 Introduction

Adolescent pregnancy has important health and social consequences, and the reproductive needs of married adolescents warrant attention in Bangladesh.[2, 3] Evidence useful for policy must distinguish the frequency of births from the proportion of adolescents who have already had a birth or are pregnant. It must also distinguish estimates among married adolescents from estimates among all adolescent girls.

Age at first cohabitation is relevant both to union formation and to the duration during which childbearing can occur. Bongaarts’s framework describes how background circumstances relate to fertility through proximate reproductive processes.[4] This provides a substantive organising framework, but does not establish which interview-time variables are valid confounders of a causal effect. Prior Bangladesh studies have documented socioeconomic and demographic associations with adolescent childbearing.[5, 6]

The 2022 BDHS provides reproductive histories and measured household and partner characteristics within a complex sampling design. Its short and long questionnaires differ in the information collected.[3] Consequently, exclusions due to questionnaire assignment should not be treated as ordinary item nonresponse. A systematic review documented incomplete design handling and model-assessment reporting in DHS/MICS analyses across countries.[1] These findings motivate transparent implementation and reporting rather than a claim that every Bangladesh study is incorrect. Current age is also important: respondents interviewed at age 15 and at age 19 have experienced different observation windows.

This study therefore has two objectives: to describe childbearing or current pregnancy among ever-married adolescents aged 15–19, and to examine conditional associations with cohabitation timing and partner characteristics among currently married long-questionnaire respondents. It is an association study, not an evaluation of a marriage law, reproductive intervention, or causal pathway.

## 2 Methods

### 2.1 Data source and design

The 2022 BDHS is a nationally representative household survey with a two-stage stratified cluster design. Selected households were interviewed and eligible ever-married women aged 15–49 were invited to individual interviews. A long questionnaire was administered in two-thirds of selected households; the remaining households received a short questionnaire covering background characteristics and reproductive history.[3] The supplied individual recode contains 30,078 women, 674 observed primary sampling units (PSUs), and 16 strata. The report’s planned PSU count and the observed recode count should not be conflated.

### 2.2 Populations and sample selection

The descriptive population comprised 2,449 ever-married respondents aged 15–19. Questionnaire assignment was identified directly using `SQTYPE`: 814 received the short questionnaire and 1,635 the long questionnaire. Among long-questionnaire respondents, 32 were formerly married and 1,603 were currently married. The partner-characteristics analysis was defined for currently married respondents because husband education and age were unavailable for the formerly married group. Two currently married respondents reported unknown husband education; the final regression sample was 1,601 (Figure 1).

**Figure 1.**
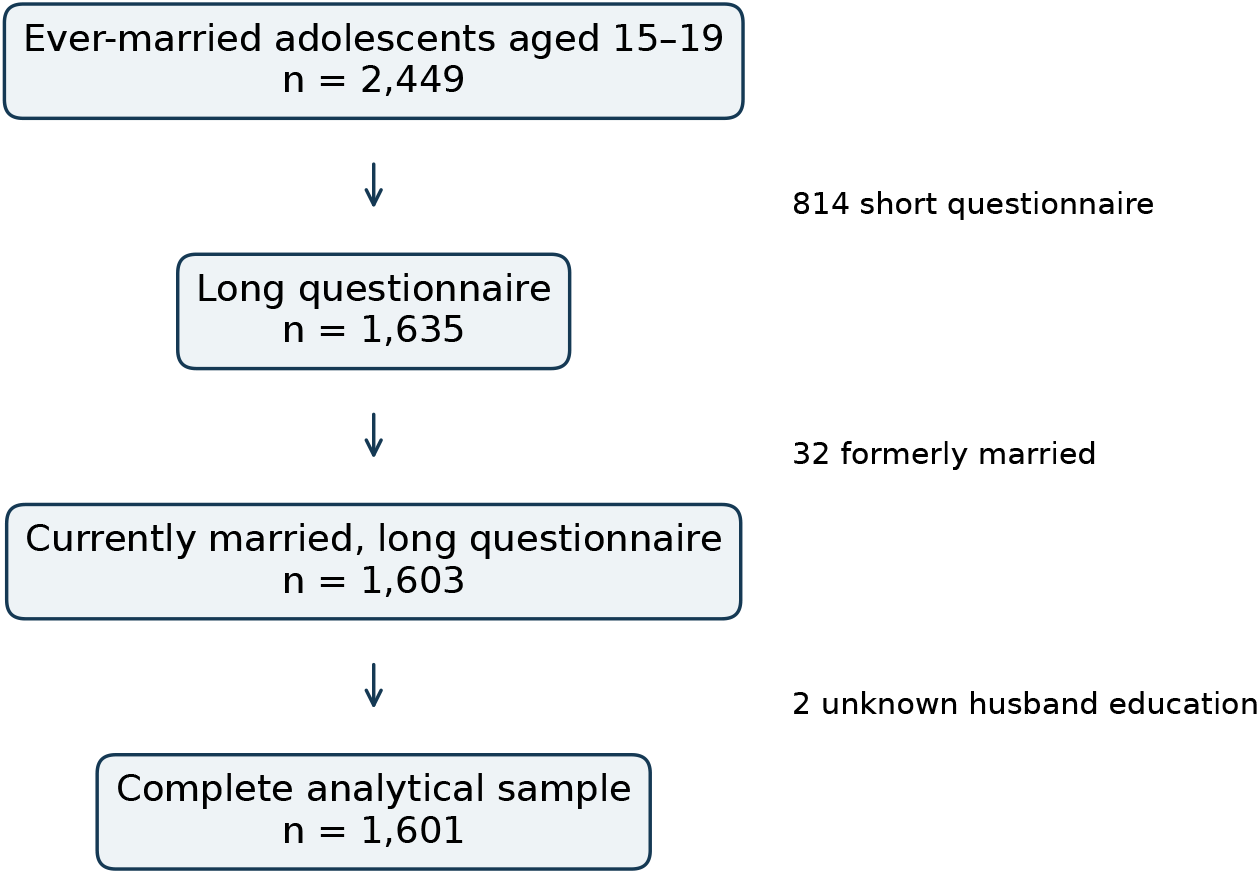
Verified sample selection. The regression concerns currently married long-questionnaire respondents; the descriptive population includes all 2,449 ever-married adolescents.

This selection is distinct from excluding 848 respondents for ordinary missing responses. No missing values were imputed. Short-questionnaire respondents remain in descriptive estimates for variables measured in both questionnaire forms. No a priori sample-size calculation was conducted; available survey records were used, and category counts and precision were examined rather than assuming adequacy from total sample size alone.

### 2.3 Outcome

The binary outcome equalled one if total children ever born (`V201`) exceeded zero or current pregnancy (V213) equalled one; otherwise it equalled zero. Both source variables were observed for the descriptive population. The pregnancy variable’s zero category denotes “no or unsure”; uncertainty is not distinguishable within that code. The recoded outcome exactly matched the original derived dataset among eligible adolescents.

The measure captures lifetime live-birth experience or current pregnancy at interview among respondents currently aged 15–19. It includes any live births before age 15. It is neither a birth rate nor a complete measure of all pregnancies, because previous pregnancies without a live birth are not included unless the respondent is currently pregnant.

### 2.4 Exposure and covariates

The main exposure was first-cohabitation age (V511), grouped as below 15, 15–17, and 18–19 years. The expanded model also included current age (five separate categories), respondent education (none, primary, secondary, higher), original household wealth quintiles (poorest to richest), husband education (none, primary, secondary, higher), signed spousal age gap (at most five, 6–10, at least 11 years), urban/rural residence and eight administrative divisions. The gap was calculated as husband age minus respondent age; four negative differences are included in the at-most-five category.

Variables were selected using the study question and plausible demographic and geographic relationships, rather than bivariate significance or stepwise selection. Current age addresses the observation window; residence and division describe geographic context. Socioeconomic and partner characteristics allow conditional descriptions but are not asserted to form a sufficient causal adjustment set. Education, marital-household wealth and location were measured at interview and may not represent premarital circumstances. Current working status was excluded from the expanded model and included in a sensitivity model because employment may follow childbearing. Confounder selection for a causal question would require additional temporal and substantive justification.[7, 8]

### 2.5 Statistical analysis

Weights were V005/1,000,000, with V021 for PSUs and V023 for strata. Analytic-domain estimates retained the full recode’s PSU/stratum structure. Descriptive outcome confidence intervals used Taylor linearisation and a logit transformation. Regression used inverse-probability-weighted logistic estimation and a stratified ultimate-cluster Taylor sandwich: PSU score totals were centred within strata, with zero contributions for PSUs outside the analytic domain. No finite population correction was applied. Regression intervals used a *t* reference distribution with 539 residual degrees of freedom for the main model, following the R survey package default for the analytic domain (581 occupied PSUs minus 16 strata minus 26 non-intercept parameters). The original full-PSU covariance structure is retained.[9]

All covariates in each specification were entered simultaneously. Odds ratios describe odds, not prevalence ratios or percentage-point changes. Reported coefficient-level *p*-values are exploratory; no omnibus bivariate test or multiplicity-adjusted confirmatory claim is made. This avoids reproducing the original script’s minimum-category-*p* screening as a factor-level test.

The analysis was executed in Python using `pyreadstat, pandas, numpy, scipy, statsmodels` and `patsy`, with explicit survey covariance calculations. Independent verification was executed using R 4.3.3 and survey 4.2. Coefficients and survey standard errors agreed to numerical precision. The residual degrees-of-freedom convention was reconciled with R and confidence intervals were updated. Applicability of the individual weight and variance approach to long-questionnaire subsampling has not yet been confirmed; regression estimates using these weights are provisional pending reconciliation with questionnaire-specific guidance.

### 2.6 Sensitivity analyses and diagnostics

We compared the historical six-predictor specification, an age-adjusted version, an age-and-geography cohabitation model, the expanded model, and an expanded model additionally including work. A duration-based timing specification replaced cohabitation groups with a cubic spline in years since first cohabitation; it addresses a different timing parameterisation, not an independently identified cohabitation effect. A robust Poisson specification was assessed but not adopted as the principal probability model.

An exploratory weighted PCA used eight binary asset items: electricity, radio, television, refrigerator, bicycle, motorcycle/scooter, car/truck and landline telephone. Codes other than zero/one were treated as ineligible for that asset construct. PCA was based on the weighted correlation matrix in the asset-complete analytical subset. PC1 was scaled to unit weighted variance and oriented towards positive refrigerator ownership. It replaced official wealth only in a sensitivity model; an official-wealth model on the same respondents was fitted for comparison. PCA did not select confounders. The official DHS wealth index is already derived from asset PCA.[10]

Completed diagnostics included convergence, linear-program checks for complete and quasi-separation, matrix rank, term-level generalized variance inflation factors (GVIFs), category counts, age/cohabitation overlap, and independent point-estimation checks. Each of the 581 occupied PSUs was deleted in turn and the model refitted to assess selected-coefficient sensitivity. A stratified ultimate-cluster bootstrap used 200 replicates, preserving the fixed model specification. These bootstrap intervals are approximate and do not incorporate unavailable lower-stage design details or a finite population correction.

Internal validation used ten repetitions of five-fold partitions of PSUs within strata. Respondents from the same PSU stayed together. Weighted AUC, Brier score, calibration intercept and slope were calculated from predictions for held-out PSUs. These are internal assessments of a fixed specification chosen after earlier results were seen. Repetition ranges are not confidence intervals and do not remove optimism from historical model selection.

An exploratory specification diagnostic added quadratic and cubic powers of the standardized fitted logit to the model and used a survey-adjusted Wald test. It is an approximate post hoc diagnostic with generated regressors, rather than a definitive omnibus goodness-of-fit test. External validation and causal-identification assumptions remain unverified.

## 3 Results

### 3.1 Outcome prevalence and sample characteristics

Among the 2,449 ever-married adolescents, weighted prevalence of the outcome was 59.7% (95% CI 57.6–61.7). Figure 2 shows age-specific descriptive estimates, reflecting different ages and observation windows. These estimates should not be generalised to never-married adolescents.

**Figure 2.**
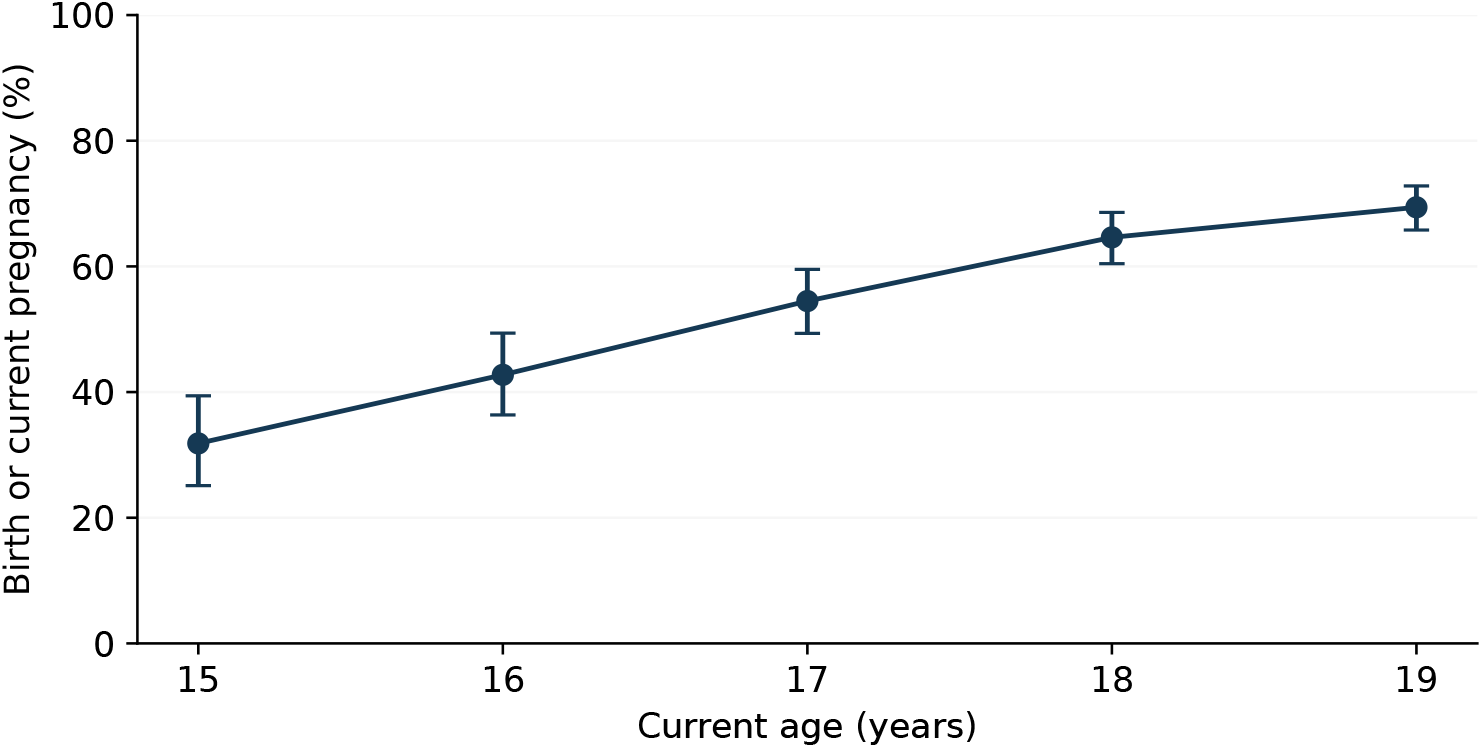
Survey-weighted prevalence of a live birth or current pregnancy by current age among ever-married adolescents (*n* = 2, 449). Bars denote 95% confidence intervals. These are cross-sectional age comparisons, not a longitudinal trajectory.

The regression sample contained 960 respondents with the outcome and 641 without it. Table 1 describes its characteristics. The no-education reference category contained only 25 respondents, including six without the outcome, which contributes to imprecision in education comparisons. Cohabitation at 18–19 was observed only among respondents currently aged 18–19; comparisons with younger current ages lack support.

**Table 1.** Characteristics of the regression sample (*n* = 1, 601).

| Characteristic | Category | Unweighted $n$ | Weighted % |
| --- | --- | --- | --- |
| Current age | 15 | 128 | 8.1 |
|  | 16 | 180 | 11.3 |
|  | 17 | 257 | 16.4 |
|  | 18 | 489 | 30.7 |
|  | 19 | 547 | 33.4 |
| Respondent education | None | 25 | 1.5 |
|  | Primary | 223 | 13.3 |
|  | Secondary | 1231 | 78.1 |
|  | Higher | 122 | 7.1 |
| Household wealth | Poorest | 316 | 19.3 |
|  | Poorer | 351 | 22.1 |
|  | Middle | 358 | 21.6 |
|  | Richer | 346 | 22.0 |
|  | Richest | 230 | 14.9 |
| First cohabitation age | Below 15 | 425 | 26.9 |
|  | 15–17 | 948 | 58.2 |
|  | 18–19 | 228 | 14.9 |
| Husband education | None | 117 | 6.9 |
|  | Primary | 429 | 26.6 |
|  | Secondary | 693 | 44.8 |
|  | Higher | 362 | 21.6 |
| Signed spousal age gap | $\leq 5$ | 528 | 31.9 |
|  | 6–10 | 702 | 43.5 |
| | $\geq 11$ | 371 | 24.5 |
| Residence | Urban | 482 | 23.6 |
|  | Rural | 1119 | 76.4 |
| Division | Barishal | 171 | 6.1 |
|  | Chattogram | 235 | 19.0 |
|  | Dhaka | 257 | 24.9 |
|  | Khulna | 223 | 12.5 |
|  | Mymensingh | 169 | 7.1 |
|  | Rajshahi | 223 | 14.1 |
|  | Rangpur | 225 | 13.1 |
|  | Sylhet | 98 | 3.2 |

### 3.2. Conditional associations

In the expanded model excluding work, cohabitation at 15–17 versus below 15 was associated with lower odds (AOR 0.170, 95% CI 0.113–0.256), as was cohabitation at 18–19 (AOR 0.026, 0.015–0.045). A gap of at least 11 versus at most five years was associated with higher odds (AOR 1.752, 1.241–2.474). Husband higher education versus none was inconclusive (AOR 0.612, 0.338–1.109); the interval is compatible with a substantial inverse association as well as little association. Current age was strongly associated with the outcome: age 19 versus 15 had an AOR of 52.855 (95% CI 28.619–97.615). Richer and richest households had lower adjusted odds than the poorest (AORs 0.633, 0.408–0.980, and 0.390, 0.238–0.637). Several division comparisons indicated higher odds than Barishal, although estimates near the null threshold should be interpreted cautiously given multiple exploratory comparisons. Respondent education, rural residence, a gap of 6–10 years, and primary or secondary husband education had intervals including one. Figure 3 displays these selected contrasts; Table 2 provides every non-intercept coefficient, avoiding selective presentation.

**Table 2.** Expanded-model conditional associations, excluding work (*n* = 1, 601).

| Contrast | AOR | 95% CI | *p* |
| --- | --- | --- | --- |
| Age: 16 | 4.569 | 2.431–8.587 | < 0.001 |
| Age: 17 | 9.179 | 5.346–15.760 | < 0.001 |
| Age: 18 | 20.590 | 11.500–36.866 | < 0.001 |
| Age: 19 | 52.855 | 28.619–97.615 | < 0.001 |
| Respondent education: Primary | 0.964 | 0.287–3.238 | 0.953 |
| Respondent education: Secondary | 0.881 | 0.257–3.021 | 0.840 |
| Respondent education: Higher | 0.930 | 0.251–3.440 | 0.913 |
| Wealth: Poorer | 0.724 | 0.483–1.085 | 0.117 |
| Wealth: Middle | 0.877 | 0.574–1.340 | 0.544 |
| Wealth: Richer | 0.633 | 0.408–0.980 | 0.041 |
| Wealth: Richest | 0.390 | 0.238–0.637 | < 0.001 |
| Cohabitation age: 15–17 | 0.170 | 0.113–0.256 | < 0.001 |
| Cohabitation age: 18–19 | 0.026 | 0.015–0.045 | < 0.001 |
| Husband education: Primary | 1.123 | 0.613–2.059 | 0.706 |
| Husband education: Secondary | 0.911 | 0.506–1.642 | 0.757 |
| Husband education: Higher | 0.612 | 0.338–1.109 | 0.105 |
| Spousal age gap: 6–10 | 0.904 | 0.682–1.197 | 0.480 |
| Spousal age gap: $\geq 11$ | 1.752 | 1.241–2.474 | 0.001 |
| Residence: Rural | 0.968 | 0.717–1.307 | 0.834 |
| Division: Chattogram | 2.093 | 1.329–3.298 | 0.001 |
| Division: Dhaka | 1.613 | 1.013–2.567 | 0.044 |
| Division: Khulna | 2.016 | 1.211–3.356 | 0.007 |
| Division: Mymensingh | 1.104 | 0.645–1.890 | 0.717 |
| Division: Rajshahi | 1.449 | 0.846–2.484 | 0.177 |
| Division: Rangpur | 1.648 | 1.002–2.712 | 0.049 |
| Division: Sylhet | 1.563 | 0.820–2.981 | 0.174 |

**Table 3.** Selected AORs (95% CIs) across specifications.

| Contrast | Historical | Age/geography | Expanded | Expanded + work |
| --- | --- | --- | --- | --- |
| Cohabitation 15–17 vs <15 | 0.409 (0.306–0.548) | 0.156 (0.105–0.234) | 0.170 (0.113–0.256) | 0.170 (0.113–0.256) |
| Cohabitation 18–19 vs <15 | 0.150 (0.101–0.225) | 0.024 (0.014–0.040) | 0.026 (0.015–0.045) | 0.026 (0.015–0.045) |
| Higher husband education vs none | 0.457 (0.270–0.774) | – | 0.612 (0.338–1.109) | 0.612 (0.338–1.109) |
| Spousal age gap $\geq 11$ vs $\leq 5$ | 1.626 (1.198–2.206) | – | 1.752 (1.241–2.474) | 1.752 (1.242–2.473) |

**Figure 3.**
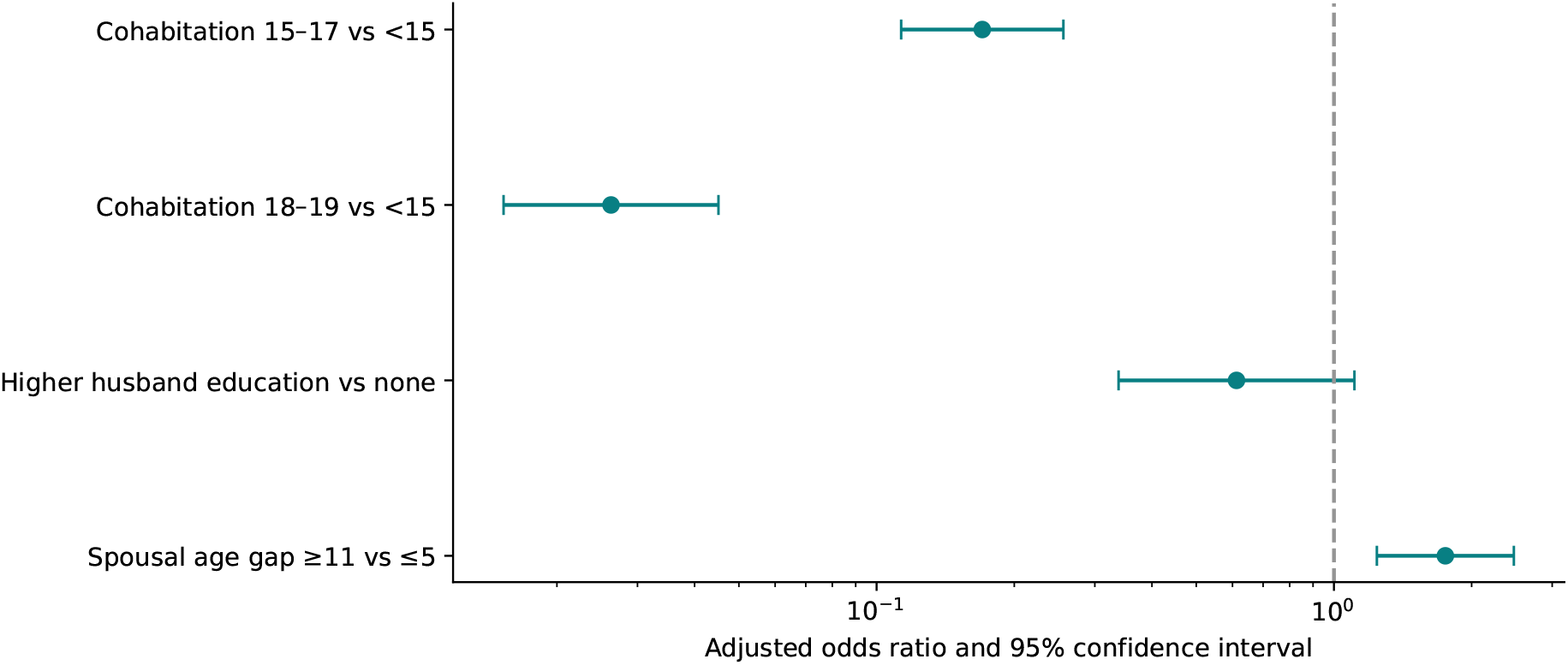
Selected expanded-model odds ratios and 95% confidence intervals (*n* = 1, 601). The dotted line denotes an odds ratio of one. All comparisons are conditional associations; the cohabitation contrasts also reflect differing time available for childbearing.

Reference categories: age 15; no respondent education; poorest wealth; cohabitation below 15; no husband education; gap at most five; urban residence; Barishal. All listed variables are included together. *p*-values are coefficient-level exploratory comparisons, not omnibus factor tests.

### 3.3 Sensitivity and model behaviour

The historical six-predictor model reproduced the original cohabitation AORs of approximately 0.409 and 0.150. Adding current age changed these to 0.168 and 0.027. Therefore, the inverse association persists but its magnitude is specification-dependent. Adjustment should not be chosen according to whether an association becomes stronger or weaker. Adding geography and retaining five wealth quintiles yielded the expanded-model estimates above. Additional work adjustment changed the selected estimates negligibly (Supplementary Table 3).

All fitted specifications converged. Apparent weighted AUC increased from approximately 0.694 in the historical model to approximately 0.811 in the expanded model including work; these are descriptive training-sample values, not evidence of external performance or absence of bias. The Poisson specification generated fitted means as high as 1.487 and was not adopted as the main probability model. The duration-spline specification had a high information-matrix condition number and near-one fitted probabilities, motivating further stability assessment rather than a claim of superiority.

The asset PCA used 1,299 of the 1,601 respondents: 302 had not-de-jure asset codes. PC1 explained 19.3% of standardized asset variation. This modest fraction, rare asset ownership and selected subset do not justify replacing the official wealth index in the main analysis. Full model estimates, asset loadings and component variance fractions are included in the reproducibility package.

### 3.4 Independent verification and internal validation

The main model had full column rank (27 of 27 parameters), converged, and showed no complete or quasi-separation within numerical tolerance. The maximum adjusted term GVIF was 1.182. Dummy-column VIFs were larger because of factor coding and the small reference group; these were not interpreted as independent evidence for removing a factor.

R reproduced the AORs with a maximum absolute difference of approximately 1.4 *×* 10^*−*11^ and survey standard errors within 2.7 *×* 10^*−*8^. The main residual degrees of freedom were 539 under the R analytic-domain convention, compared with 632 in the earlier implementation. The correction slightly widened intervals without changing the selected conclusions.

Across exact single-PSU deletions, the maximum change in the selected cohabitation, higher-husband-education and large-age-gap coefficients was 0.286 survey standard errors. All deletion fits and the 200 bootstrap fits had small final score residuals. Bootstrap percentile intervals retained lower odds for the cohabitation groups and higher odds for the large age gap; higher husband education remained inconclusive.

Apparent weighted AUC was 0.811 and mean held-out-PSU AUC was 0.793 (partition range 0.789–0.798). Apparent and mean held-out Brier scores were 0.171 and 0.180, respectively. Mean calibration slope was 0.885 (range 0.868–0.910) and intercept was 0.034, suggesting some optimism and predictions that were too extreme. Calibration deciles from the first partition are descriptive and shown in Figure 4.

**Figure 4.**
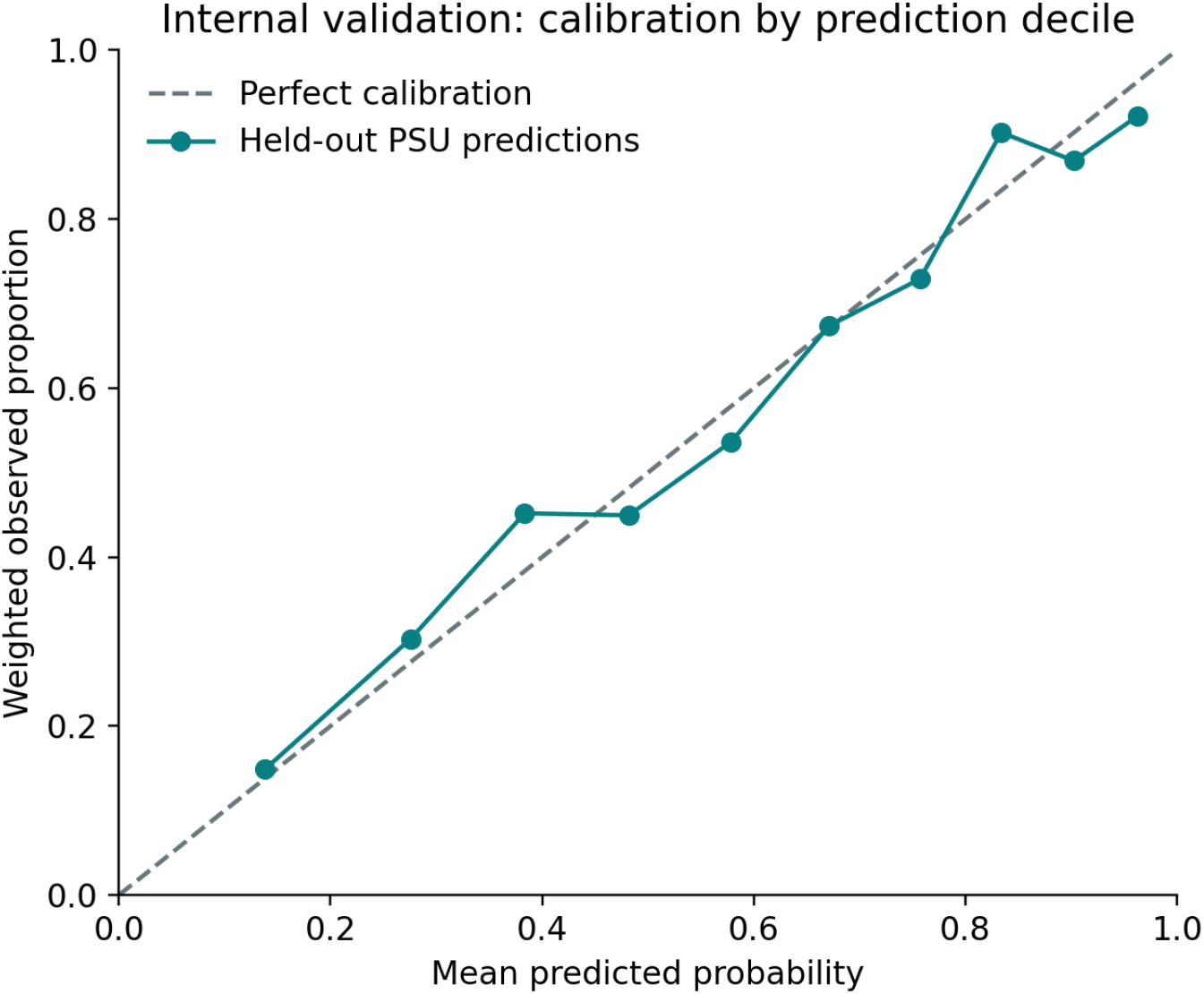
Weighted observed outcome proportions versus mean held-out-PSU predicted probabilities by decile in the first five-fold partition. The diagonal denotes ideal agreement. Points are descriptive and have no uncertainty intervals.

The exploratory fitted-logit polynomial test yielded *F* = 3.913 and *p ≈* 0.0205, indicating possible specification inadequacy. This signal warrants substantive examination of timing parameterizations and restricted-overlap comparisons. It does not identify a particular omitted mechanism, and models should not be selected solely to eliminate a significant diagnostic.

## 4 Discussion

This analysis distinguishes the descriptive ever-married population from the currently married population with partner information. The main reproducible pattern is lower odds of a live birth or current pregnancy among those beginning cohabitation later. A larger signed spousal age gap was also associated with higher odds. In contrast, the original statistically conclusive higher-husband-education result was not retained under the expanded adjustment specification.

### 4.1 Timing and interpretation

Cohabitation age and current age jointly determine the time available for childbearing after union formation. Comparing an adolescent who began cohabitation years earlier with one who began recently is consequently not equivalent to estimating an intervention effect. Age adjustment addresses an important observation-window difference but does not eliminate duration-related interpretation, residual confounding or selection into marriage. Earlier Bangladesh findings provide context for the direction of the observed pattern, not validation of a causal mechanism.[5, 6]

The partner age-gap finding may motivate research on reproductive expectations, relationship circumstances and access to services. Those pathways were not directly tested. Likewise, husband education should not be interpreted as proven irrelevant because its confidence interval crosses one. Wider intervals and changes across specifications indicate uncertainty.

### 4.2 Implications for Bangladesh

The descriptive findings motivate assessment of reproductive-health service needs among ever-married adolescents; the present outcome does not itself measure unmet need or preferences. Service planning can consider age, marital circumstances and access to appropriate antenatal and reproductive-health care. Existing external evidence and policy commitments may support preventing child marriage and supporting adolescent autonomy,[2] but this survey model does not quantify the benefits of a specific programme, legislative change, or partner-focused intervention.

Geographic estimates can identify questions for local assessment, but cross-sectional differences should not be used alone to rank programme effectiveness. Evaluating a policy requires a defined intervention, an appropriate comparison, pre-intervention information and an identification strategy. The present model also does not estimate socioeconomic total effects: some adjusted characteristics may be consequences or intermediates.

### 4.3 Strengths and limitations

Strengths include verification against the raw recode, explicit distinction between questionnaire exclusions and unknown responses, current-age adjustment, retention of the full PSU/stratum structure, complete coefficient reporting and transparent sensitivity specifications. No unsupported imputation reassurance is retained.

Important limitations remain. First, this is cross-sectional and the outcome is cumulative through different interview ages. Second, covariates measured at interview cannot be assumed to precede cohabitation or childbearing; this permits reverse causation and inappropriate causal adjustment. Third, restricting partner analyses to currently married adolescents can select a population different from all ever-married or all adolescent women. Fourth, questionnaire-specific weighting remains unresolved, despite independent reproduction of the provisional design-based estimates in R. Internal calibration suggests some overfitting and an exploratory specification test indicates possible misspecification, so model adequacy cannot be regarded as established. Fifth, self-reported ages and dates may be affected by recall and age heaping. Sixth, sparse education categories and age/cohabitation structural zeros limit some comparisons. Finally, model revisions were developed after reviewing results; confirmatory language and selective significance claims are inappropriate.

The pregnancy coding combines “no” with “unsure”, and the composite does not include previous non-live-birth pregnancies. A future timing analysis should separate pregnancy from completed live-birth events and validate dates, censoring and births before union. More advanced estimators cannot compensate for unavailable premarital confounders or lack of a suitable target population.

## 5 Conclusion

Among currently married adolescents aged 15–19 with long-questionnaire information, later first cohabitation was associated with lower odds of having had a live birth or being currently pregnant, while a larger spousal age gap was associated with higher odds. Current-age adjustment substantially affected the cohabitation estimates. Findings describe conditional patterns and motivate service-needs assessment; they do not establish causal effects or predict the impact of a marriage policy.

## Data Availability

The data analyzed in this study are available from The DHS Program upon registration and approval of a data access request. The authors are not permitted to redistribute the dataset under The DHS Program data-use conditions.

https://dhsprogram.com/methodology/survey/survey-display-584.cfm

## Declarations

### Ethics

This is a secondary analysis without new recruitment. The original BDHS obtained ethical review and participant consent as documented in its report.[3] No new participants were recruited or contacted for this analysis. No claim of a separate institutional exemption or approval is made here.

### Data availability

The original BDHS data are available through the DHS Program subject to registration and access conditions. Raw respondent data are not redistributed with the analysis package. The reproducibility materials comprise analysis scripts, derived aggregate tables and figure-generation files; these exclude raw respondent data.

### AI assistance

ChatGPT and Codex (OpenAI) were used as assistive tools for statistical review, code drafting, figure preparation, language editing and manuscript revision. Md. Ikramul Haq Khan personally reviewed and checked the analytical workflow, code, outputs, interpretations and manuscript drafts. The authors reviewed and approved this revision and retain responsibility for the accuracy, integrity and conclusions of the work. AI assistance supported the research process and did not replace human scientific judgment or responsibility. AI tools are not listed as authors. These human checks do not imply that the analytical limitations reported in this manuscript have been resolved.

### Author approval

All authors agreed to this revision and the authorship listed on the title page. Md. Ikramul Haq Khan personally checked the analysis and manuscript preparation.

## A Supplementary model comparisons

Historical: six original predictors including work, without age or geography. Age/geography: cohabitation groups, current age, residence and division. Expanded: age, education, five wealth quintiles, cohabitation groups, husband education, gap, residence and division. Expanded + work adds working status. Dashes denote variables not entered, not missing estimates. All models use the same 1,601 respondents. Differences cannot be interpreted solely as removal of confounding; logistic odds ratios are conditional and specification-dependent.

## B Reproducibility and remaining analytical questions

The analysis used executed Python and independent R implementations. The reproducibility materials described for the analysis include aggregate coefficient tables, category counts, overlap tables, PCA loadings and component variances. The editable LaTeX package supplied with this manuscript contains the manuscript source, tables and figures; it does not contain respondent-level data or constitute the complete analytical code archive. The optional ggplot2 script was not used to generate the manuscript figures.

The authors approved the revised question, model set and conclusions. Long-questionnaire weighting guidance still requires confirmation. The specification signal and timing comparisons with restricted overlap also warrant further investigation. If analytical specifications or weights change, estimates and uncertainty calculations must be recomputed and checked against the independent implementation. Internal validation does not establish external performance or causal identification.

