## Supplemental Table S1 for "Cohabitation timing and childbearing or current pregnancy among married adolescents in Bangladesh: a complex-survey analysis of the 2022 BDHS" (from another version of this preprint: version 2, 2026-07-24)

**Supplementary Material**

**S1 Table.** Survey-weighted multivariable logistic regression of factors associated with adolescent fertility, multiple imputation sensitivity analysis (n = 2,449).

| **Variable** | **Category** | **AOR (95% CI)** | **p-value** |
| --- | --- | --- | --- |
| **Education** | No education | (Ref.) | – |
|  | Primary | 0.69 (0.30, 1.63) | 0.403 |
|  | Secondary | 0.73 (0.32, 1.67) | 0.453 |
|  | Higher | 0.95 (0.39, 2.35) | 0.917 |
| **Household wealth** | Poor | (Ref.) | – |
|  | Middle | 0.85 (0.66, 1.08) | 0.185 |
|  | Rich | 0.76 (0.60, 0.96) | 0.020 |
| **Age at first cohabitation (years)** | <15 | (Ref.) | – |
|  | 15–17 | 0.45 (0.33, 0.60) | <0.001 |
|  | 18–19 | 0.15 (0.10, 0.23) | <0.001 |
| **Husband's education** | No education | (Ref.) | – |
|  | Primary | 0.95 (0.57, 1.58) | 0.849 |
|  | Secondary | 0.70 (0.44, 1.13) | 0.146 |
|  | Higher | 0.45 (0.27, 0.75) | 0.003 |
| **Spousal age gap (years)** | ≤ 5 | (Ref.) | – |
|  | 6–10 | 0.92 (0.70, 1.20) | 0.523 |
|  | ≥ 11 | 1.66 (1.21, 2.27) | 0.002 |
| **Respondent currently working** | No | (Ref.) | – |
|  | Yes | 1.23 (0.83, 1.82) | 0.299 |

*AOR, adjusted odds ratio; CI, confidence interval; Ref., reference category. Missing data on the explanatory variables were addressed using multiple imputation by chained equations across all 2,449 eligible ever-married adolescent women aged 15–19 years. Twenty imputed datasets were generated; the survey design (sampling weights, primary sampling units, and stratification) was applied to each imputed dataset, and estimates were pooled using Rubin's rules. The outcome and survey design variables were not imputed. The same model specification and reference categories as the complete-case analysis (Table 3) were used. Estimates with a 95% CI excluding 1.00 (p < 0.05) are statistically significant.*

The pooled estimates from the multiply imputed datasets were consistent with the complete-case analysis reported in the main text. Age at first cohabitation remained the strongest factor, with markedly lower odds of adolescent fertility among women who first cohabited at 15–17 years (AOR = 0.45, 95% CI: 0.33–0.60) and 18–19 years (AOR = 0.15, 95% CI: 0.10–0.23) relative to those who cohabited before age 15 years. Higher partner education (AOR = 0.45, 95% CI: 0.27–0.75) remained protective, and a spousal age gap of 11 years or more remained associated with higher odds (AOR = 1.66, 95% CI: 1.21–2.27). Household wealth showed a significant inverse association in the imputed analysis (rich vs poor: AOR = 0.76, 95% CI: 0.60–0.96). Respondent educational attainment and current working status were not independently associated with adolescent fertility. The direction and magnitude of associations were largely unchanged from the complete-case model, indicating that the findings were robust to the handling of missing data.
